## Supplementary analyses for "The R = 1 threshold can misclassify epidemic stability"

### Supplementary Note 1: Additional empirical example (Texas, USA)

In the main text, we demonstrated the complications from assessing epidemic stability using standard estimators of transmissibility in **Fig 5** for COVID-19 Veneto, Italy. Here we consider an additional example as a sensitivity or validation test and confirm that scenarios we exposed in the main text were not special or rarely occurring in real data. Our analysis looks at COVID-19 cases in the top 24 counties (by total cases, these account for 75% of all cases in the state) that compose Texas in the USA. This data is sourced from https://github.com/nytimes/covid-19-data and we compute the standard reproduction number $R$, the risk-averse reproduction number $E$ and the probability of either statistic (denoted $X$) being above 1. See main text for more details and the formulae for computing these estimates.

The results below corroborate the main text and suggest the relative ease of finding scenarios where $R=1$ is an unreliable measure of stability due to infections being aggregated over many heterogeneous groups. In **Fig S1** we highlight two periods of discrepancy between $R$ and $E$. In the first, the estimated R=1 because the total infections are relatively stable. However, we see that many individual counties are growing, which is reflected in $E>1$ and eventually $R$ also grows above 1. The second period is approximately stable as E=1 and the incidence of many counties is stable or weakly growing. There is a decline in the incidence of the county with the most infections, which $R$ overly weights ($E$ balances this against the weak growth). The result is an extremely confident $R<1$ estimate. The slowly growing counties eventually dominate the spread and $E$ tracks this uptick in transmissibility notably earlier than $R$.

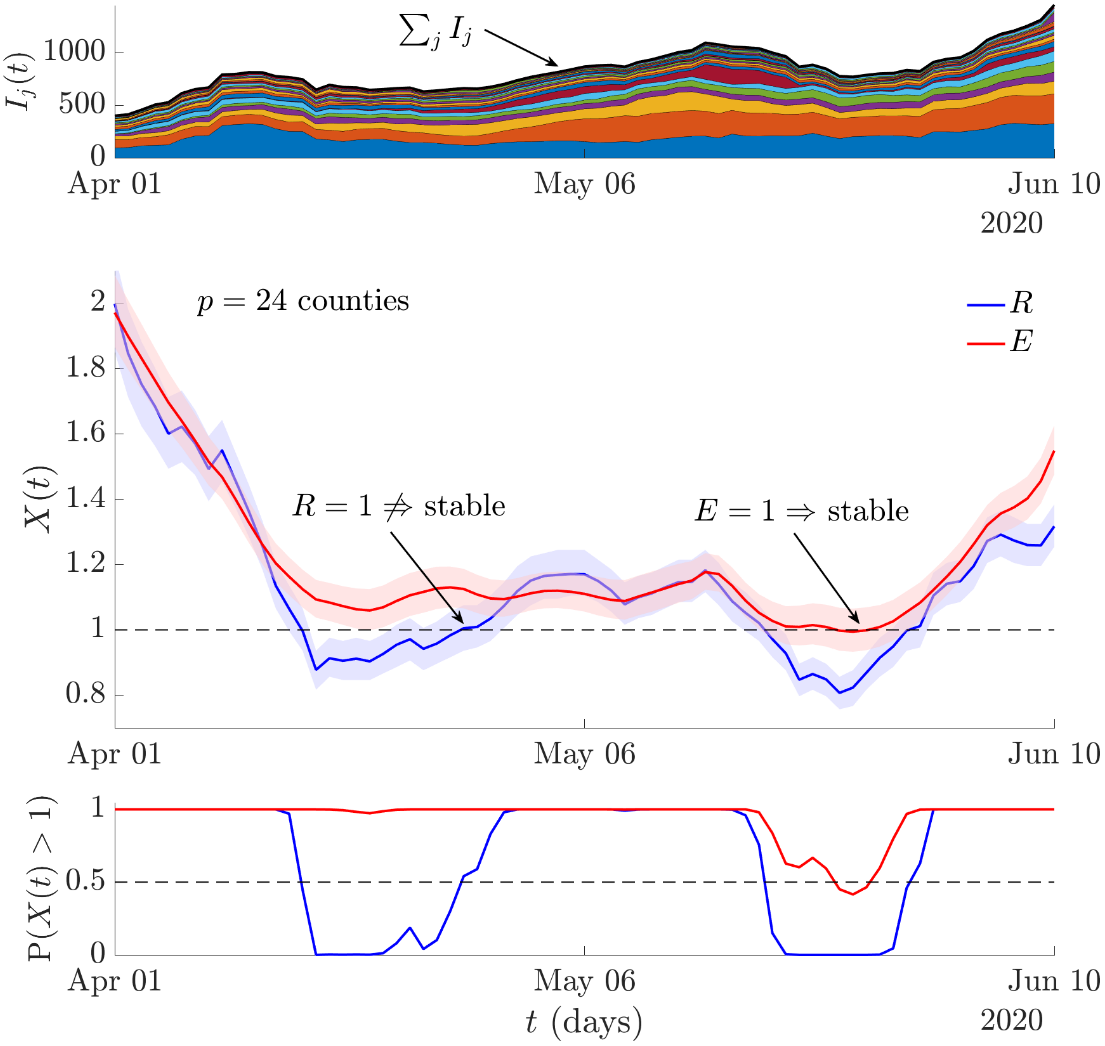


**Fig S1**: **False stability thresholds for COVID-19 in Texas, USA.** We examine COVID-19 dynamics across $p = 24$ groups, which are counties with the most cases in the state of Texas. Data are the incidence of new cases derived from the New York Times repository (see link in text). The cases are smoothed with a 7-day moving filter to limit weekend effects and negative values are set to 0 before smoothing to remove artefacts. The top panel stacks these incidence curves $I_j(t)$ by counties in order of their total cases (so the first area plot corresponds to the county with most cases) reaching the eventual total infection time series (shown in black). We estimate the standard $R(t)$ (blue) and the risk averse $E(t)$ reproduction number (red) using EpiFilter in the middle panel and compute the probability that these statistics (broadly denoted $X(t)$) are above 1 in the bottom panel. We observe two periods of divergence between *R* and *E*. The first has a false positive signal of stability (*R=1*, *E>1*), while in the second *R* confidently signals decline, while *E* suggests critical stability. See text for discussion of these periods.

### Supplementary Note 2: Epidemic thresholds for interacting groups

In the main text, we commented on the suitability of *R* and *E* in cases where real time response is not imperative and where there is access to rich contact-network data. Here we investigate when these statistics and particularly *E* may still work reliably given interactions are known and significant. We also expose how the true contact-data aware threshold *T* depends on the accuracy of that data by analysing how it markedly changes over 4 scenarios below.

We consider a heterogeneous renewal model (see Methods) broadly defined below with $p_{kj}$ as the probability of an infection generated in location *k* migrating into location *j*.

$$I_j(t) = m_j(t) + p_{jj}R_j\Lambda_j + \sum_{k\neq j} p_{kj}R_k\Lambda_k. \quad \text{(i)}$$

Here $\Lambda_j = \int_0^t G(t-x)I_j(x)\,\mathrm{d}x$ and the $m_j(t) = 0$ except in the location where the epidemic starts, then $m_j(t) = \delta(t)$. We assume the same generation time distribution across locations for notational convenience and use deterministic equation forms (though including Poisson noise is straightforward). As the migration probabilities are conserved $\sum_j p_{kj} = 1$.

An important point is immediate from (i). If we look at the total infections $\sum_j I_j(t) = I(t)$ and collect our sums of migration terms, then we obtain $I(t) = \delta(t) + \sum_j R_j\Lambda_j$. Comparing to the commonly assumed global model $I(t) = \delta(t) + \Lambda R$, we find that under any migration scheme the standard *R* is unchanged from the formulation that neglects interactions. Consequently, in these cases this popular *R* may still be interpreted as a meaningful baseline.

We can take Laplace transforms of (i) to get (ii) below. This yields a matrix of transfer functions (see the Methods in the main text for more information about transfer functions).

$$I_j(s) = m_j(s) + p_{jj}R_jG(s)I_j(s) + \textstyle\sum_{k\neq j} p_{kj}R_kG(s)I_k(s). \quad \text{(ii)}$$

To better explore the diversity of transmission possibilities emerging from interaction, we solve the equations for 2 demes or locations only. However, similar analyses can be applied to larger group or deme numbers. We denote $p_{11}$ as $p_1$ and note $p_{12} = 1 - p_1$. Symmetrical arguments apply for $p_{22}$ and $p_{21}$ leading to the characteristic polynomial $\Delta(s)$ for this system in (iii).

$$\Delta(s) = \big(1 - p_1R_1G(s)\big)\big(1 - p_2R_2G(s)\big) - (1-p_1)(1-p_2)R_1R_2G^2(s). \quad \text{(iii)}$$

Poles of the renewal model of (i) solve $\Delta(s) = 0$. The dominant pole is the overall growth rate.

For a critical dominant pole (zero growth), we require that parameters in (iii) satisfy $\Delta(s = 0) = 0$. This yields the condition (iv) i.e., if our $p_j$ and $R_j$ satisfy this relation then we are certain that the true epidemic threshold *T* is at 1 and the overall epidemic is critically stable.

$$p_1 R_1 + p_2 R_2 - 1 = (p_1 + p_2 - 1) R_1 R_2. \quad \text{(iv)}$$

As (iv) uses all possible information, we do not expect *E* to perform as well as the resulting metric obtained from this condition. However, we can show the value of *E* on key conditions below. We define the threshold of stability by rearranging (iv) into a form $T = 1$.

1. Antisymmetric migration: $p_1 = 1 - p_2$. In this case, the threshold stability satisfies $p_1 R_1 + p_2 R_2 = 1$, which is a weighted mean of the local reproduction numbers. Under any choice of $p_1$ we recover a version of **Fig 1** from the main text, where *E* is better than *R*. However, the fully informed $T = p_1 R_1 + p_2 R_2$ will always perform best but requires us knowing the migration probabilities as well as local reproduction numbers accurately.

2. Symmetric migration: $p_1 = p_2 = \rho$. Substituting this into (iv) we get $\rho(R_1 + R_2) - 1 = (2\rho - 1) R_1 R_2$. If we take small deviations $(\delta, \epsilon)$ about 1, so that $R_1 \approx 1 + \delta$ and $R_2 \approx 1 + \epsilon$, then $T = 1$ requires $(\delta + \epsilon)(1 - \rho) \approx 0$ while $E = 1$ requires $0.5(\delta + \epsilon) \approx 0$. Hence, when $\rho \neq 1$, *E* locally approximates the correct threshold condition. If $\rho = 0.5$, the approximation becomes exact. If instead $\rho = 1$, we recover the non-interacting case.

3. Dominant (unidirectional) migration: $p_2 = 1$. Critical stability requires $p_1 R_1 + R_2 - 1 = 1 + p_1 R_1 R_2$. This is the same as the non-interacting case of the main text, but the local reproduction numbers are now $p_1 R_1$ and $R_2$. If $p_1 = 1$, they are $p_2 R_2$ and $R_1$. Adjusting the definition of *E* to these new local reproduction numbers recovers our results from the main text, but it is clear even unidirectional interactions change our notion of local spread.

4. Symmetric transmissibility: $R_1 = R_2 = \mu$. Now (iv) becomes $(p_1 + p_2 - 1)\mu^2 - (p_1 + p_2)\mu + 1 = 0$. This quadratic is solved by $T = \mu = 1$. Computing $E = \sum_j R_j^2 / \sum_j R_j = \mu$ we find *E* is precisely the true stability threshold for any migration probability.

Consequently, *E=1* can in several instances be a good approximator for overall stability. While less accurate than *T*, *E* requires far less information, which makes it apt for real-time analyses. *T* and mobility or contact-matrix based methods are advantageous for retrospective analyses from high resolution data. However, if the $p_{kj}$ are not well known or change due to unexpected dynamics, computing *T* is difficult and biases can emerge (e.g., 1-4 are notably diverse).
